# The synergistic impact of serology and nucleic acid test to enhance blood transfusion safety: a retrospective observational study among blood donors at tertiary care hospital in Pakistan

**DOI:** 10.1101/2025.07.30.25332420

**Authors:** Uzma Ata, Arisha Sohail, Samra Waheed, Salman Ali, Uzma Bukhari

## Abstract

**Objective:** This study aimed to evaluate the impact of combined use of chemiluminescent immunoassay (CLIA) and nucleic acid amplification testing (NAAT) to improve transfusion transmitted Hepatitis B Virus (HBV), Hepatitis C virus (HCV) and Human Immunodeficiency Virus-1 (HIV-1) among blood donors in Pakistan.

**Design:** Retrospective, single center observational.

**Setting:** Regional blood center at tertiary care hospital in urban Pakistan.

**Participants:** All adults of 18 years or above who were eligible to donate blood after meeting the pre-donation screening criteria during the study period (n = 26,778).

**Outcome Measures:** Primary outcome measure was to estimate the incremental yield of HBV, HCV, and HIV-1 infections using combined CLIA+NAAT compared to CLIA-alone per 100,000 donations. Secondary outcome was to calculate inter-test agreement and discordance between CLIA and NAAT methods.

**Results:** Among 26,778 donors, the combined CLIA+NAAT testing detected a total of 2,423.6 viral infections per 100,000 donations, NAAT alone contributed 739.7/100,000 of these. For HBV, NAAT uniquely detected 489/100,000 additional cases missed by CLIA; the combined detection rate was 1,561/100,000. For HCV, NAAT-only yield was lower (247/100,000), with total detection of 825.3/100,000. HIV-1 was rare in the donor pool; incremental NAAT yield was 3.7/100,000, with a combined detection rate of 37.3/100,000. Agreement between tests was substantial for HBV (κ = 0.63) and moderate to fair for HCV (κ = 0.47) and HIV-1 (κ = 0.40). The discordant cases detected by NAAT alone for HBV, HCV and HIV-1 were 183, 431 and 37 respectively. McNemar’s test showed statistically significant differences (p < 0.001) across all markers, with large effect sizes for HIV-1 (0.92, 95% CI: 0.80–1.00) and HCV (0.69, 95% CI: 0.65–0.73).

**Conclusion:** Integrating CLIA with NAAT enhanced the detection of HBV, including occult and window period infections, and refined estimates of active HCV and HIV-1 infections which significantly improved blood transfusion safety.

**STRENGTHS AND LIMITATIONS OF THIS STUDY:**

- The selection bias was reduced by including all consecutive eligible blood donors during study period.
- The information bias was minimized through uniform, fully automated, and validated protocols for combined CLIA and NAAT testing of blood donors.
- The single center retrospective study design restricts generalizability and causal inference.
- The false positives/negatives and genetic variability were not confirmed.

## INTRODUCTION

Blood transfusion plays vital role in saving millions of human lives worldwide but also carries a public health risk due to the potential transfer of infectious diseases, particularly where the prevalence of transfusion-transmitted infections (TTIs) is notably high1. Pakistan faces significant blood transfusion safety challenges particularly with the endemic prevalence of hepatitis B virus (HBV) and hepatitis C virus (HCV) which currently impact around 7.4% of the general population^1^.

The blood transfusion regulation is fragmented in Pakistan requiring more robust regulation^2^. There is a need of more than 5 million blood donations every year in Pakistan, but current annual rate of blood donations is limited to around 2.3 million donations of which about 1.9 million are family or replacement donors^3^. This high volume of replacement donors presents safety challenges, as replacement donors demonstrate continuously increased rates of TTIs than voluntary donors ^4,5^. A systematic review of 33 studies from 2010-2020 in Pakistan reported 2.04% HBV, 2.44% HCV, 0.038% HIV, 1.1% syphilis and 0.11% malaria frequency showing a significant infection rate among voluntary non-remunerated donations (VNRDs) at 0.48% than replacement donors at 4.15% ^1^.

Since Hepatitis B and C are endemic in Pakistan, the present challenges in transfusion safety practices include inadequate TTI testing due to only serology testing The blood screening is performed using ELISA, rapid diagnostic tests, and nucleic acid testing in Pakistan which has improved the blood transfusion safety but the inconsistent screening practices present constant challenges. The Chemiluminescent Immunoassay (CLIA) based blood screening shows increased sensitivity and specificity compared to conventional blood screening methods, however, it can yield false positive results, particularly in populations with increased prevalence of cross-reactive antibodies^6^. Whereas, the Nucleic Acid Amplification Testing (NAAT) directly detect viral nucleic acids by augmenting specific regions of viral DNA or RNA which significantly reduces the diagnostic window period offering higher sensitivity and specificity than serological methods^7–9^.

The combined application of CLIA and NAAT offers several complementary benefits as CLIA allows rapid screening with high throughput and NAAT adds superior sensitivity during the window period when serological tests may yield negative results^10^. Emerging evidence in Pakistan indicates that combining NAAT with serological screening provides more accurate risk estimates and lower infection rates by decreasing the residual risk of transfusion-transmissible viral infections^11–14^. This combined methodology improves the detection rates of both, acute infections through NAAT and established infections through CLIA. However, the fragmented screening practices and coverage strongly urges the need of actionable data to assess the incremental yield of combined CLIA and NAAT screening in HBV and HCV endemic areas to improve transfusion safety and inform national policy. Therefore, we aim to assess the improvement in blood screening by using combined CLIA and NAAT methods among blood donors at regional blood center in a tertiary care hospital to meet the gaps in transfusion safety and effective TTI screening strategies in Pakistan.

## METHODS

This retrospective study was carried at the Regional Blood Center of tertiary care hospital in urban Pakistan from May 2023 to December 2024. A total of 26,788 blood donors were screened and their data was recorded. No sample size calculation was performed, as the study included all eligible donations during the predefined timeframe. All voluntary and replacement blood donors who visited the Regional Blood Centre during this time were eligible to participate in the study. All adult donors aged 18 years or above went through a standardized pre-donation screening process that involved a medical history review and a physical examination, following regulatory guidelines. Blood samples from all qualified donors were included in the analysis. The standard protocol followed at the study site is shown in figure 1.

**Figure 1:**
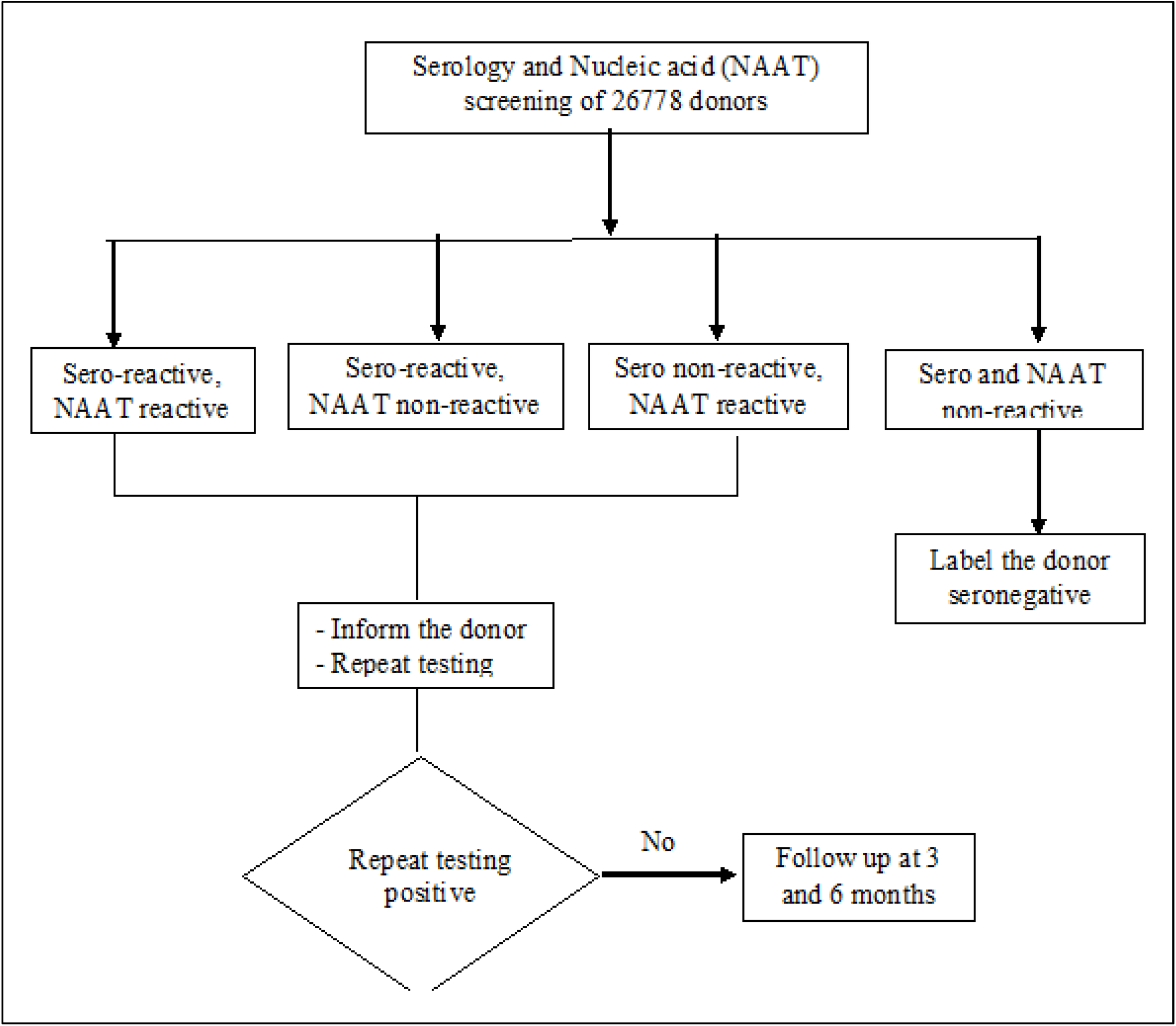
Flow chart for blood donor screening

### INSERT FIGURE 1

For the purpose of serological testing (CLIA), the Cobas e 402 analyzer (Roche Diagnostics) was utilized to identify HBsAg, anti-HCV, and anti-HIV-1. NAAT was conducted on the Cobas 5800 analyzer (Roche Diagnostics) to test for HBV DNA, HCV RNA, and HIV RNA, employing an individual donor testing (ID-NAT) format without sample pooling. All procedures were carried out in compliance with the manufacturers’ guidelines and relevant quality control standards. Consistency in testing methods was maintained throughout all donor samples. To mitigate selection bias, all consecutive eligible donors were included during the specified study period. Information bias was reduced by utilizing automated analyzers that adhered to validated protocols. The duplicate testing of initially reactive samples further minimized the potential measurement error. All initially reactive donors were contacted and referred to the appropriate authority after proper counseling.

Data were compiled in Microsoft Excel and analyzed using IBM SPSS Statistics, Version 26.0 (Armonk, NY: IBM Corp). The missing data was handled by replacing with mean values. Descriptive statistics summarized donor demographics, with continuous variables reported as means ± standard deviations and categorical variables presented as frequencies and percentages. The agreement between two test modalities for HBV, HCV, and HIV-1 was assessed using Cohen’s kappa coefficient (κ). The interpretation of agreement strength was: ≤ 0.20 (slight), 0.21– 0.40 (fair), 0.41–0.60 (moderate), 0.61–0.80 (substantial), and > 0.80 (almost perfect). To assess discordance between the two tests, McNemar’s test was used with a significance level of p < 0.05. Discordant cases (NAAT > CLIA) were recorded for each marker. Effect size estimates and 95% confidence intervals (CIs) were calculated. To assess the impact of testing on transfusion safety, yield rates per 100,000 donations were calculated for NAAT-only and combined CLIA+NAAT approaches. These rates were used to estimate potentially interdicted infections by multiplying reactive cases by three, reflecting the average number of blood components from a single whole blood donation. All tests adhered to the approved study protocol (IRB-3526/DUHS/EXEMPTION/2024/182) and the Declaration of Helsinki principles.

## RESULTS

A total of 26,778 blood donors were screened between May 2023 and December 2024 after passing blood donation eligibility criteria using a questionnaire and physical examination. The demographics of the study cohort of donors are summarized in table 1. The donor pool is predominantly male (99.5%). Almost all donations are exchanged (98%) with a small voluntary fraction (2%). Nearly every unit is collected in-house (98%); only a minor share comes from external blood drives (2%).

**Table 1:** Demographics of blood donors (*n* = 26,778)

| Characteristic |  | Value |
| --- | --- | --- |
| Age, years (Mean ± S.D) | | 28.6 ± 7.0 |
| Gender n (%) | Male | 26664 (99.5) |
|  | Female | 114 (0.42) |
| Donation type n (%) | Exchange | 26194 (98) |
|  | Voluntary | 582 (2) |
| Draw site n (%) | In-House | 26294 (98%) |
|  | External Unit | 486 (2%) |

About 3.23 % of all donations were positive for at least one transfusion-transmissible infection (TTI) by both CLIA and NAAT. Among these, the seroprevalence of anti-HCV is the most frequent at 2.19%, followed by HBsAg at 1.76% and anti-HIV-1 at 0.17%. However, after screening each blood donor using both CLIA and NAAT simultaneously, the frequency of HBV increased to 2.3%, while the frequencies of HCV and HIV-1 decreased to 0.9% and 0.03%, respectively (figure 2).

**Figure 2:**
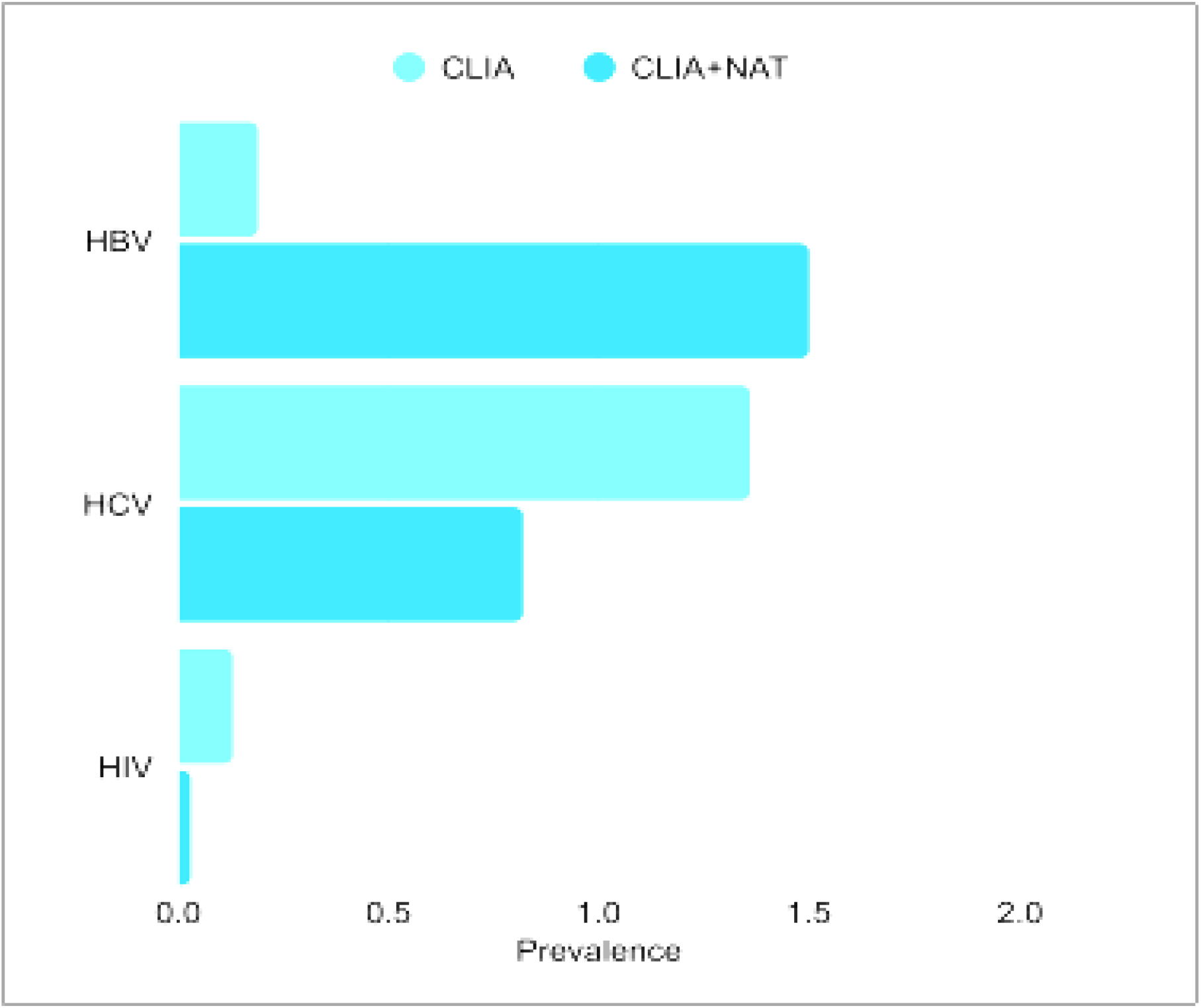
Comparison of CLIA only and CLIA +NAAT blood donor screening prevalence of HBV, HCV, and HIV-1 among 26778 blood donors.

### INSERT FIGURE 2

The CLIA sensitivity was highest for HIV-1 (0.97), moderate for HCV (0.89), and lowest for HBV (0.78), reflecting variable effectiveness across infections (**Supplementary table 1 and 2**). **TABLE 2** shows the inter-test agreement as Cohen’s kappa (_κ_), the number of cases missed by CLIA, whereas NAAT identified, and the results of McNemar’s paired test. HBV demonstrates substantial agreement (_κ_ = 0.63) with a moderate level of discordance (183 cases, p < 0.001, effect size = 0.43 [95% CI: 0.35 – 0.51]), whereas HCV (_κ_ = 0.47) and HIV-1 (_κ_ = 0.40) show only moderate to fair agreement along with significantly greater discordance: HCV with 431 cases (effect size = 0.69 [0.65 – 0.73]) and HIV-1 with 37 cases (effect size = 0.92 [0.80 – 1.00]), both p < 0.001. **FIGURE 3** shows the chi-square (χ^2^) statistic from McNemar’s test for each viral marker showing markedly raised χ ^2^ for HCV (**Supplementary Table 3**).

**Table 2:** Agreement and discrepancies between CLIA and NAAT in HBV, HCV, and HIV□1 screening among blood donors (n= 26,778).

| **Viral marker** | **Kappa ($\kappa$)** | **Discordant Cases (NAT > CLIA)** | **McNemar's Test p-value** | **Effect Size** | **95% Confidence Interval** |
| --- | --- | --- | --- | --- | --- |
| **HBV** | 0.63 | 183 | < 0.001 | 0.43 | 0.35 – 0.51 |
| **HCV** | 0.47 | 431 | < 0.001 | 0.69 | 0.65 – 0.73 |
| **HIV-1** | 0.4 | 37 | < 0.001 | 0.92 | 0.80 – 1.00 |

**Figure 3.**
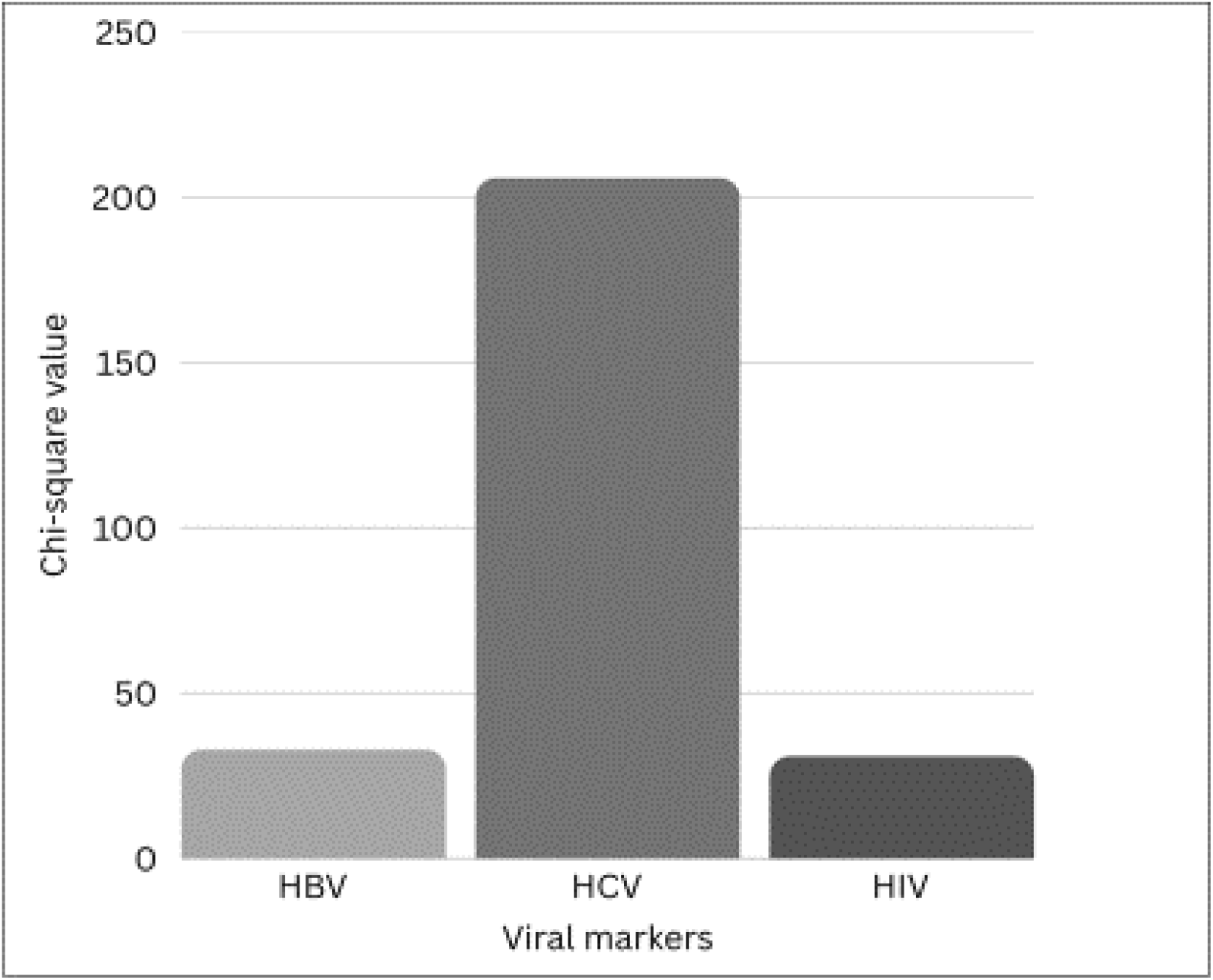
Comparison of the inter-test discordance between CLIA and NAAT in HBV, HCV, and HIV-1 screening among blood donors based on McNemar’s chi-square values.

### INSERT FIGURE 3

Among all donations tested, a total of 739.7 viral infections per 100,000 donations were uniquely detected by NAAT, while the combined NAAT and CLIA yield rate reached 2,423.6 per 100,000 donations (Table 3).

**Table 3:** Combined and individual yields of HBV, HCV, and HIV-1 by CLIA and NAAT testing among blood donors (n=26,778).

| **Viral Marker** | **CLIA reactive/NAAT non-reactive n (%)** | **CLIA non-reactive/NAAT reactive n (%)** | **CLIA reactive/NAAT reactive n (%)** | **NAAT Yield Rate (per 10<sup>5</sup> donations)** | **NAAT + CLIA Yield Rate (per 10<sup>5</sup> donations)** |
| --- | --- | --- | --- | --- | --- |
| HBV | 52 (0.19) | 131 (0.48) | 418 (1.55) | 489 | 1561 |
| HCV | 365 (1.35) | 66 (0.24) | 221 (0.82) | 247 | 825.3 |
| HIV-1 | 36 (0.13) | 1 (0.003) | 10 (0.03) | 3.7 | 37.3 |
| Combined (Total) | 451(1.68) | 196 (0.73) | 649 (2.42) | 739.7 | 2,423.6 |

HBV had the highest extra yield from NAAT, finding infections not caught by CLIA, with 489 out of 100,000 donations. This means about 1 extra HBV-infected unit was found for every 204 donations. When using both CLIA and NAT together, they detected 1,561 out of 100,000 donations, reducing the extra infected units to about 1 for every 64 donations. HCV showed a different pattern, with 364 detections by CLIA only and 65 by NAT only. The NAAT yield is 247 per 100,000 or 1 in every 405 donations. This rate is approximately half that of HBV. When both methods are used together, HCV-infected donations drop to approximately 37 per 100,000, or 1 in every 2,700 donations. The HIV-1 rate was very low in this group of donors. There were 35 cases detected only by CLIA, one case was detected only by NAT, and 10 cases were detected by both methods. NAAT detects approximately 3.7 cases per 100,000 donations, which is approximately 1 in 27,000 donations. The combined detection rate is approximately 37 per 100,000, similar to that of HCV, but mostly due to serology. Combining CLIA with NAAT testing improved detection sensitivity by more than three times for HBV and HCV, and over ten times for HIV-1, resulting in approximately 3.3 times overall enhancement (**Supplementary table 4**). The potential number of prevented infections based on component separation into three red cells, plasma, and platelets was 3,888 from each donation.

## DISCUSSION

The present study found an overall TTI prevalence of 3.23% among the blood donors, which reflects moderate endemicity consistent with previous reports in Pakistan^1,15^ (1). However, a study at a tertiary hospital in Islamabad reported TTI prevalence of 2.0% during 2018-2019 which is lower than the prevalence documented in some other studies during the same period^1^. The disparity can be connected to infection rate trends, advancements in screening strategies, or demographic changes in the populations studied. This emphasizes the variation in TTI distribution rates through various regions and blood safety strategies.

The current study shows that the pathogen-specific detection patterns with combined testing differ as compared to only serological testing. The CLIA based HBV detection rate was 1.76% which increased to 2.3% when NAAT was performed on same samples. On contrary, the CLIA based HCV detection rate paradoxically decreased from 2.19% to 0.90% when NAAT was performed on same samples. A systematic review of 106 studies (1998–2010) indicated a mean HBV prevalence of 3.93% among healthy blood donors in Pakistan, with a general carrier rate of 3–5%^16^. However, recent studies show lower HBV rates in specific regions, like a 2021 study in Khyber Pakhtunkhwa reported 0.86% HBV prevalence among 6,311 blood donors, study conducted in Karachi (2019–2020) reported 1.4% to 1.6% HBV frequency; and a 2022 study in Faisalabad found 2.8% HBV prevalence^14,17,18^. The majority of these investigations have employed serological tests such as ELISA and CLIA to detect HBsAg. On the other hand, the HCV prevalence shows greater variability from 0.3% to 7% with a pooled national estimate of 2.7%; however, NAAT-based studies consistently reported lower rates of active infection than serology alone ^19,20^. Our study found low CLIA based HIV prevalence of 0.17% which was further reduced to 0.03% when NAAT was performed on same samples. This finding is consistent with the national data, possibly influenced by cultural and behavioral factors, such as circumcision and social stigma, which may limit both transmission and reporting^14,21,22^. Despite the low prevalence, the more than tenfold increase in detection sensitivity for HIV with combined CLIA and NAAT testing highlights the critical role of NAAT in minimizing residual transfusion risk, especially given the severe consequences of transfusion-transmitted HIV. The window periods for HIV vary by pathogen as it can extend to 40 days from the last risk behavior and the CLIA screening can identify infections as early as 20 days^23^. However, CLIA may be associated with false positive results, particularly in populations with high prevalence of cross-reactive antibodies ^24^. Recent studies have established refined window periods using NAAT screening which are 5.1 to 6.6 days for HIV, 2.7 to 3.5 days for HCV, and 16.6 to 22.4 days for HCV infections^25,26^. These dramatically shortened window periods compared to serological testing represent a major advancement in blood safety.

Our study shows a moderate to fair concordance between CLIA and NAAT screening in the serological identification of HCV and HIV infections within the Pakistani donor population. Cohen’s kappa coefficients of 0.47 for HCV and 0.40 for HIV revealed limited agreement between the two modalities. Moreover, these observed discrepancies were statistically verified through significant McNemar’s test (p < 0.001) which demonstrated non-random discordance that signifies clinical and operational concern. These findings highlight continued limitations in assay performance due to differences in analytical sensitivity, specificity thresholds, and the genetic heterogeneity of circulating viral strains in the region. The discordance supports similar studies, which collectively advocate for combined screening protocols to improve transfusion safety and mitigate residual risk in blood banking^11,21,27^. Our study demonstrated a considerable level of agreement between CLIA and NAAT when screening for HBV with a kappa value of 0.63. However, a significant number of HBV cases were only detected by NAAT. These HBV cases are likely window-period donations which serological tests often miss, making NAAT superior for detecting infections early and preventing unsafe blood transfusions.

We also estimated the highest additional NAAT yield of 489 per million donations for HBV infection. When we combined CLIA and NAAT methods, the overall detection rate was refined to 1,561 cases per 100,000 donation which means about 1 out of every 64 donors tested positive for HBV which reduced the risk of missing an HBV-infected unit further. However, the HCV infection shows lower incremental NAAT detection of 247 per million donations as compared to HBV detection. The possible reasons could be the persistence of anti-HCV antibodies after viral clearance, the shorter window period for antibody development and high serological assay sensitivity. Some CLIA positives may be false due to cross-reactivity, inflating CLIA-only numbers. Therefore, adding NAAT screening of blood donor samples confirms active infection and reduces apparent prevalence. The HIV-1 prevalence in the study blood donor pool was very low, resulting in lower NAAT yield of about 1 per 27,000 and a combined detection rate of approximately 37 per 100,000 donations. More CLIA positive donors and fewer NAAT-only positives indicate that the blood donor pool has established infections with detectable antibodies, while window-period infections are rare due to low incidence and effective donor screening.

When a unit of whole blood is separated into three components (red blood cells, plasma, and platelets) one infected donation can pose a serious risk to multiple patients^28^. This study demonstrates that through combined testing, we can prevent up to 3,888 possible infections by isolating components from just one donation. Further, the 3.3-fold overall enhancement in TTI detection sensitivity in the study donor pool must be weighed against resource constraints in Pakistan’s healthcare system^29^. This emphasizes the critical threat posed by undetected transfusion-transmitted infections (TTIs) and the urgent need to address them. This risk amplification factor makes the incremental detection sensitivity provided by combined testing economically justifiable despite higher costs.

## LIMITATIONS

The methodological aspects of this study have few limitations. The study’s retrospective design limits causal inference, and the absence of confirmatory testing details for discordant results raises questions about false positive/negative rates. The pooling strategy for NAAT testing could affect sensitivity, particularly for low-viral-load samples. Additionally, the lack of genotyping data prevents assessment of whether discordant results reflect viral genetic diversity affecting assay performance

## CONCLUSION

This study demonstrates that the integration of chemiluminescent immunoassay (CLIA) and nucleic acid amplification testing (NAAT) does not provide uniform advantages across all transfusion-transmissible infections (TTIs). The notable enhancement in HBV detection when NAAT is incorporated underscores the presence of HBV occult and infections within window-period that would otherwise remain undetected. Conversely, a decrease in HCV detection with the combined testing suggests that NAAT can refine the estimation of active infections, thereby improving donor screening and preventing the unnecessary deferral of donors with resolved infections, particularly in resource-limited settings.

## FUTURE WORK

Future research should focus on longitudinal monitoring of TTI trends, cost-effectiveness analyses of combined testing strategies, and investigation into viral genotypic diversity affecting assay performance. Strengthening reporting of transfusion reactions and donor adverse events will be essential to evaluate the real-world impact of enhanced screening protocols and guide continuous quality improvement. Successful implementation requires training and quality assurance deficits identified in national surveys. Studies have shown that technical validations are conducted in only 48% of public sector blood centers, highlighting significant quality assurance gaps, which is further complicated by the lack of mandatory accreditation systems in Pakistan.

## Data availability statement

Data are available upon reasonable request.

## Ethics statements

Patient consent for publication

Not applicable.

## Ethics approval

This study involves human participants and was exempted by Dow University of Health Sciences Ethics Committee (IRB-3526/DUHS/EXEMPTION/2024/182) because the electronic health record was used. Each participants gave informed consent to participate in the study at the time of sreening..

## Acknowledgments

We are grateful to the technical staff for their assistance in obtaining the study data at regional blood center.

## Footnotes

Contributors UA was responsible for conceptualisation, data acquisition, interpretation, supervision, review and editing. SW and SA were responsible for data acquisition and interpretation, study coordination and editing. AS was responsible for writing original draft, data analysis, interpretation. UB was responsible for the overall content as the guarantor. All the authors approved the final manuscript as submitted and agree to be accountable for all aspects of the work.

## Funding

This research received no specific grant from any funding agency in the public, commercial or not-for-profit sectors’

## Competing interests

None declared.

## Patient and public involvement

Patients and/or the public were not involved in the design, or conduct, or reporting, or dissemination plans of this research.

## Notes

### Competing Interest Statement

The authors have declared no competing interest.

### Funding Statement

This study did not receive any funding

### Author Declarations

This study involves human participants and was approved by Dow University of Health Sciences Ethics Committee (IRB-3526/DUHS/EXEMPTION/2024/182). Participants gave informed consent to participate in the study before taking part.

## REFERENCES

1. Ehsan H, Wahab A, Shafqat MA, et al. A Systematic Review of Transfusion-Transmissible Infections Among Blood Donors and Associated Safety Challenges in Pakistan. J Blood Med. 2020;Volume 11:405–420. doi:10.2147/JBM.S277541

2. Zaheer HA, Waheed U. Blood safety system reforms in Pakistan. Blood Transfus. Published online 2014. doi:10.2450/2014.0253-13

3. Khan H. WHO says Pakistan receives less than half of 5 million blood donations it needs annually. Accessed July 1, 2025. https://arab.news/phygx

4. Mremi A, Yahaya JJ, Nyindo M, Mollel E. Transfusion-Transmitted Infections and associated risk factors at the Northern Zone Blood Transfusion Center in Tanzania: A study of blood donors between 2017 and 2019. Gregori L, ed. PLOS ONE. 2021;16(3):e0249061. doi:10.1371/journal.pone.0249061

5. PuertoLJMeredith S, Singogo E, Chagomerana M, et al. Systematic review of prevalence and risk factors of transfusion transmissible infections among blood donors, and blood safety improvements in Southern Africa. Transfus Med. 2023;33(5):355–371. doi:10.1111/tme.12988

6. Shahin D, Aly R, Ghannam M, et al. A comparative analysis between NAT and chemiluminescence in detection of transfusion transmitted viruses in two main university blood transfusion centers. Sci Rep. 2025;15(1):20109. doi:10.1038/s41598-025-03506-6

7. Wang H, Li G, Zhao J, Li Y, Ai Y. An Overview of Nucleic Acid Testing for the Novel Coronavirus SARS-CoV-2. Front Med. 2021;7:571709. doi:10.3389/fmed.2020.571709

8. Zilouchian H, Faqah O, Kabir MA, et al. Current and Future Diagnostics for Hepatitis C Virus Infection. Chemosensors. 2025;13(2):31. doi:10.3390/chemosensors13020031

9. Sharma P, Batheja G, Verma HN, Seth P. Development of a novel molecular assay based on conventional PCR liquid hybridization and ELISA (NAT-ELISA) for the detection of HIV, HCV, and HBV in blood donors. Indian J Med Microbiol. 2022;40(1):122–126. doi:10.1016/j.ijmmb.2021.12.014

10. Cuccorese M, Roli L, Trenti T. Comparative assessment of Maglumi SARS-CoV-2 antigen test and nucleic acid amplification test. Biochim Clin. 2023;(2-2023). doi:10.19186/BC_2023.016

11. Ali SM, Raza N, Irfan M, Mohammad MF, Kazmi FH, Fatima Z. Effectiveness of Using Nucleic Acid Amplification Test to Screen Blood Donors for Hepatitis B, Hepatitis C, and HIV: A Tertiary Care Hospital Experience From Pakistan. Cureus. Published online 25 January 2023. doi:10.7759/cureus.34216

12. Niazi SK, Bhatti FA, Salamat N, Ghani E, Tayyab M. Impact of nucleic acid amplification test on screening of blood donors in Northern Pakistan. Transfusion (Paris). 2015;55(7):1803–1811. doi:10.1111/trf.13017

13. Sultan S, Nasir MI, Rafiq S, Baig MA, Akbani S, Irfan SM. Multiplex real-time RT-PCR assay for transfusion transmitted viruses in sero-negative allogeneic blood donors: an experience from Southern Pakistan. Malays J Pathol. 2017;39(2):149–154.

14. Irshad R, Farooq U, Shah T, Gul A, Badshah A. HEPATITIS B, C AND HIV EXPOSURE ON NAT AND CLIA BLOOD EXAMINATION METHODS AT REGIONAL BLOOD CENTRE ABBOTTABAD. J Ayub Med Coll Abbottabad. 2023;35(2). doi:10.55519/JAMC-02-11600

15. Sabir N, Ghafoor T, Fatima S, Lodhi R, Mehmood A, Zaman G. Prevalence and Association of Transfusion-Transmissible Infections with Age of Blood Donors: A Regional Transfusion Centre Study in Northern Pakistan. J Coll Physicians Surg--Pak JCPSP. 2023;33(9):978–982. doi:10.29271/jcpsp.2023.09.978

16. Ali M, Idrees M, Ali L, et al. Hepatitis B virus in Pakistan: A systematic review of prevalence, risk factors, awareness status and genotypes. Virol J. 2011;8(1). doi:10.1186/1743-422x-8-102

17. Ahmed S, Méndez RY, Naveed S, et al. Assessment of hepatitis-related knowledge, attitudes, and practices on quality of life with the moderating role of internalized stigma among hepatitis B-positive patients in Pakistan. Health Psychol Behav Med. 2023;11(1). doi:10.1080/21642850.2023.2192782

18. Kashif Raza S, Bajwa H, Javaid H, Anwar R, Hashim M, Saleem K. Prevalence of Transmissible Infectious Diseases among Healthy Blood Donors in Faisalabad, Pakistan: Transmissible Infectious Diseases among Healthy Blood Donors. Pak J Health Sci. Published online 31 March 2023:142-146. doi:10.54393/pjhs.v4i03.544

19. Kouser S, Qadir H, Ahmad M, et al. Nucleic Acid Amplification Testing for Human Immunodeficiency Virus, Hepatitis B Virus, and Hepatitis C Virus in Blood Donors at a Tertiary Care Hospital Blood Bank. Cureus. Published online 17 January 2025. doi:10.7759/cureus.77571

20. Waheed U, Khokhar O, Saba N, et al. The National Burden of Hepatitis C Among Blood Donors in Pakistan: A Systematic Review and Meta-Analysis. 2024/12/01. 20. doi:10.48036/apims.v20iSuppl.%202.1261

21. Irshad R, Umer Farooq, Ihsan Ullah, Abeera Farooq, Tahir Shah, Adeel Gul. EVALUATION and MEASURE OF ACCURACY OF ICT, CLIA and NAT FOR THE DETECTION OF HEPATITIS B, C and HIV IN THE BLOOD DONORS ABSTRACT. J Ayub Med Coll Abbottabad. 2023;35(4):654–657. doi:10.55519/JAMC-04-12757

22. Salman Y, Shaeen SK, Butt MS, Vohra LI, Hashmi T. HIV in Pakistan: Challenges, efforts and recommendations. Ann Med Surg 2012. 2022;84:104797. doi:10.1016/j.amsu.2022.104797

23. Ghani E, Rathore MA, Khan SA. Trends in human immunodeficiency virus seroprevalence in blood donors in northern Pakistan. Public Health. 2016;131:71–74. doi:10.1016/j.puhe.2015.10.010

24. Tan SS, Chew KL, Saw S, Jureen R, Sethi S. Cross-reactivity of SARS-CoV-2 with HIV chemiluminescent assay leading to false-positive results. J Clin Pathol. 2021;74(9):614–614. doi:10.1136/jclinpath-2020-206942

25. Dutch M, Cheng A, Kiely P, Seed C. Revised nucleic acid test window periods: Applications and limitations in organ donation practice. Transpl Infect Dis. 2024;26(1):e14180. doi:10.1111/tid.14180

26. Dutch M, Cheng A, Kiely P, Seed C. 311.6: Procleix® Ultrio Elite®Assay: Revised nucleic acid test window periods and applications in organ donation. Transplantation. 2023;107(10S1):74-74. doi:10.1097/01.tp.0000993416.32044.07

27. Pandey HC, Varghese M, Rana A, Kumar R, Jain P. Residual risk estimates of transfusion transmissible hepatitis B, hepatitis C and human immunodeficiency virus using nucleic acid testing yield/window period model in an Indian setting. Transfus Med. 2022;32(6):492–498. doi:10.1111/tme.12923

28. Rathore M, Naeem M, Javed A, Raja M. Bacterial contamination of platelets and red blood cell concentrates: A regional transfusion center study in Pakistan. Asian J Transfus Sci. 2022;0(0):0. doi:10.4103/ajts.ajts_129_20

29. Bui TI, Brown AP, Brown M, et al. Comparison of a dual antibody and antigen HCV immunoassay to standard of care algorithmic testing. Hayden R, ed. J Clin Microbiol. 2024;62(10):e00832–24. doi:10.1128/jcm.00832-24

