## supplementary file for "The synergistic impact of serology and nucleic acid test to enhance blood transfusion safety: a retrospective observational study among blood donors at tertiary care hospital in Pakistan"

**Supplementary Table 1. Contingency Analysis showing NAAT yield donations for HCV, HBV and HIV-1** (n=26,778).

| **HCV** | **NAT +** | **NAT –** |
| --- | --- | --- |
| **CLIA +** | 221 | 365 |
| **CLIA –** | 66 | 26126 |
| **HBV** | **NAT +** | **NAT –** |
| **CLIA +** | 418 | 52 |
| **CLIA –** | 131 | 26176 |
| **HIV** | **NAT +** | **NAT –** |
| **CLIA +** | 10 | 36 |
| **CLIA –** | 1 | 26731 |

**Supplementary Table 2. CLIA sensitivity (paired test) for HBV, HCV and HIV-1** (n=26,778).

| **Viral marker** | **CLIA sensitivity*** |
| --- | --- |
| HBV | 0.78 |
| HCV | 0.89 |
| HIV | 0.97 |

*Sensitivity CLIA = (a + b) / (a + b + c)

**Supplementary Table 3: McNemar’s test comparing paired diagnostic outcomes between CLIA and NAT assays for HBV, HIV, and HCV detection** (n=26,778).

| **Virus** | **CLIA+/NAT-** | **CLIA-/NAT+** | **χ² (1, N)** | **χ² Value** | **p-value** | **95% CI** |
| --- | --- | --- | --- | --- | --- | --- |
| HBV | 52 | 131 | (1, N=183) | 33.24 | < .001 | 0.220 to 0.355 |
| HIV | 36 | 1 | (1, N=37) | 31.24 | < .001 | 0.858 to 0.999 |
| HCV | 365 | 66 | (1, N=431) | ≈ 206.0 | < .001 | 0.809 to 0.880 |

**Supplementary Table 4: NAAT and Combined NAAT+CLIA Yield showing Sensitivity Improvement Factor for TTI Detection Using Combined NAAT + CLIA** (n=26,778).

| **Virus/TTI** | **NAAT Yield Rate (per 100,000)** | **Combined Yield Rate (NAAT + CLIA)** | **Improvement Factor** |
| --- | --- | --- | --- |
| HBV | 489.0 | 1,561.0 | 3.19 |
| HCV | 247.0 | 825.3 | 3.34 |
| HIV-1 | 3.7 | 37.3 | 10.08 |
| Combined TTIs | 739.7 | 2,423.6 | 3.28 |
